# Field evaluation of a rapid antigen test (Panbio™ COVID-19 Ag Rapid Test Device) for the diagnosis of COVID-19 in primary healthcare centers

**DOI:** 10.1101/2020.10.16.20213850

**Authors:** Eliseo Albert, Ignacio Torres, Felipe Bueno, Dixie Huntley, Estefanía Molla, Miguel Ángel Fernández-Fuentes, Mireia Martínez, Sandrine Poujois, Lorena Forqué, Arantxa Valdivia, Carlos Solano de la Asunción, Josep Ferrer, Javier Colomina, David Navarro

## Abstract

We evaluated the Panbio™ COVID-19 AG Rapid Test Device (RAD) for the diagnosis of COVID-19 in symptomatic patients attended in primary healthcare centers (n=412). Overall specificity and sensitivity of RAD was 100% and 79.6%, respectively, taking RT-PCR as the reference. SARS-CoV-2 could not be cultured from specimens yielding RT-PCR+/RAD- results.

## Introduction

Rapid detection, effective isolation of symptomatic cases and systematic tracing of close contacts are paramount to blunt community spread of SARS-CoV-2 infection. Nowadays, reverse□transcriptase polymerase chain reaction (RT□PCR) is the diagnostic “gold standard” for COVID-19 [1]; nevertheless, specialized instrument and expertise are required to conduct RT-PCR assays. In addition, supply shortages of RT-PCR reagents have been encountered by many countries. Rapid antigen detection (RAD) immunoassays are particularly suited for point of care testing (POC), as they can be easily performed and interpreted without equipment, are low cost, and improve the turn-around time for results. Moreover, results obtained by a recently launched antigen assay appeared to correlate better with patient infectiousness than those returned by RT-PCR [2]. In this field study, we evaluated the Panbio™ COVID-19 AG Rapid Test Device (Abbott Diagnostic GmbH, Jena, Germany), a lateral flow immunochromatographic assay targeting SARS-CoV-2 nucleoprotein in nasopharyngheal (NP) specimens for diagnosis of COVID-19 in symptomatic patients attended in primary healthcare centers.

## Material and methods

### Patients

This prospective study included 412 patients (median age, 31 years; range, 1-91; 58% females), of whom 327 were adults (median, 36 years; range, 17-91) and 85 pediatrics (≤16 years old; median, 11 years; range, 1-16), with clinical suspicion of COVID-19 attended in primary care centers (n=8) of the Health Department Clínico-Malvarrosa in Valencia. Only patients with compatible signs or symptoms appearing within the prior week (0-7 days) were recruited. The study was conducted between September 2^nd^ and October 7^th^ 2020. The study was approved by the Hospital Clínico Universitario (HCU) INCLIVA Ethics Committee.

### SARS-CoV-2 testing

Trained nurses at each participating center collected two NP per patient using flocked swabs following appropriate safety precautions, one of which (provided by the manufacturer) was used for RAD and the other was placed in 3□mL of universal transport medium-UTM-(Becton Dickinson, Sparks, MD) and delivered to the Microbiology Service of HCU for RT-PCR testing. RAD was performed immediately after sampling following the instructions of the manufacturer (reading at 15 min.). RT-PCRs were carried out within 24 h. of specimen collection with the TaqPath COVID-19 Combo Kit (Thermo Fisher Scientific, Massachusetts, USA), which targets SARS-CoV-2 ORF1ab, N and S genes. RNA was extracted using the Applied Biosystems™ MagMAX™ Viral/Pathogen II Nucleic Acid Isolation Kits coupled with Thermo Scientific™ KingFisher Flex automated instrument (Thermo Fisher Scientific). The AMPLIRUN® TOTAL SARS-COV-2 Control (Vircell S.A:, Granada, Spain-) was used as the reference material for SARS-CoV-2 RNA load quantitation (in copies/ml, considering RT-PCR C_Ts_ for the N gene: the linear regression equation was: Y = - 0.31*X + 13.77; R^2^ = 9.89).

### SARS-CoV-2 cell culture

Samples collected in UTM were stored at −80°C for up to 2 weeks before being processed for culture. Vero E6 cells, maintained in Modified Eagles Medium (MEM) supplemented with 5% Fetal Bovine Serum (FBS), 1% penicillin/ streptomycin, 0.5 µg/mL Amphotericin B and 1% L-glutamine, were seeded into 96 well plates (Thermo Fisher Scientific) at 10^5^ cells/ml and inoculated in triplicate with patient samples (100 µl of a 1:1 dilution in MEM 2% FBS supplemented with antibiotics). Cultures were incubated at 37°C with 5% CO_2_ for 4 days. Blind subcultures were performed at 48 h and incubated for another 4 days. Cytopathic effect (CPE) was evaluated daily and recorded. The presence of SARS-CoV-2 was confirmed by RT-PCR (performed at time 0, and days 2 and 4).

### Statistical analyses

RAD was evaluated for its sensitivity and specificity with the associated 95% confidence intervals (CI) using RT-PCR as the reference. Negative predictive value (NPV) and positive predictive value (PPV) were calculated for prevalences of SARS-CoV-2 infection of 5% and 10%, according to that in our Health Department within the study period. Agreement between RAD and RT-PCR was assessed using Cohen’s Kappa (κ) statistics. Differences between medians were compared using the Mann-Whitney U-test. Receiver operating characteristic curves were built to determine SARS-CoV-2 RT-PCR cycle threshold (C_T_) and RNA loads best discriminating between RT-PCR+/RAD+ or RAD-specimens. Two-sided *P*-values <0.05 were considered significant. Statistical analyses were performed using SPSS version 25.0 (SPSS, Chicago, IL, USA).

## Results

Out of 412 patients, 43 tested positive by RT-PCR and RAD (10.4%) and 358 (86.9%) negative by both methods. Discordant results (RT-PCR+/RAD-) were noticed in 11 patients (2.7%). Characteristics of these patients are shown in Supplementary Table 1. There were no RT-PCR-/RAD+ specimens. Concordance between both methods was good (κ, 0.87; 95% CI, 0.79-0.94). Accordingly, overall specificity and sensitivity of RAD was 100% (95% CI, 98.7-100%), and 79.6% (95% CI, 67.-88.8%), respectively. Sensitivity slightly increased (80.4%; 95% CI, 66.8-89.3%) in patients with clinical courses shorter than 5 days (Figure 1A).

**Figure 1.**
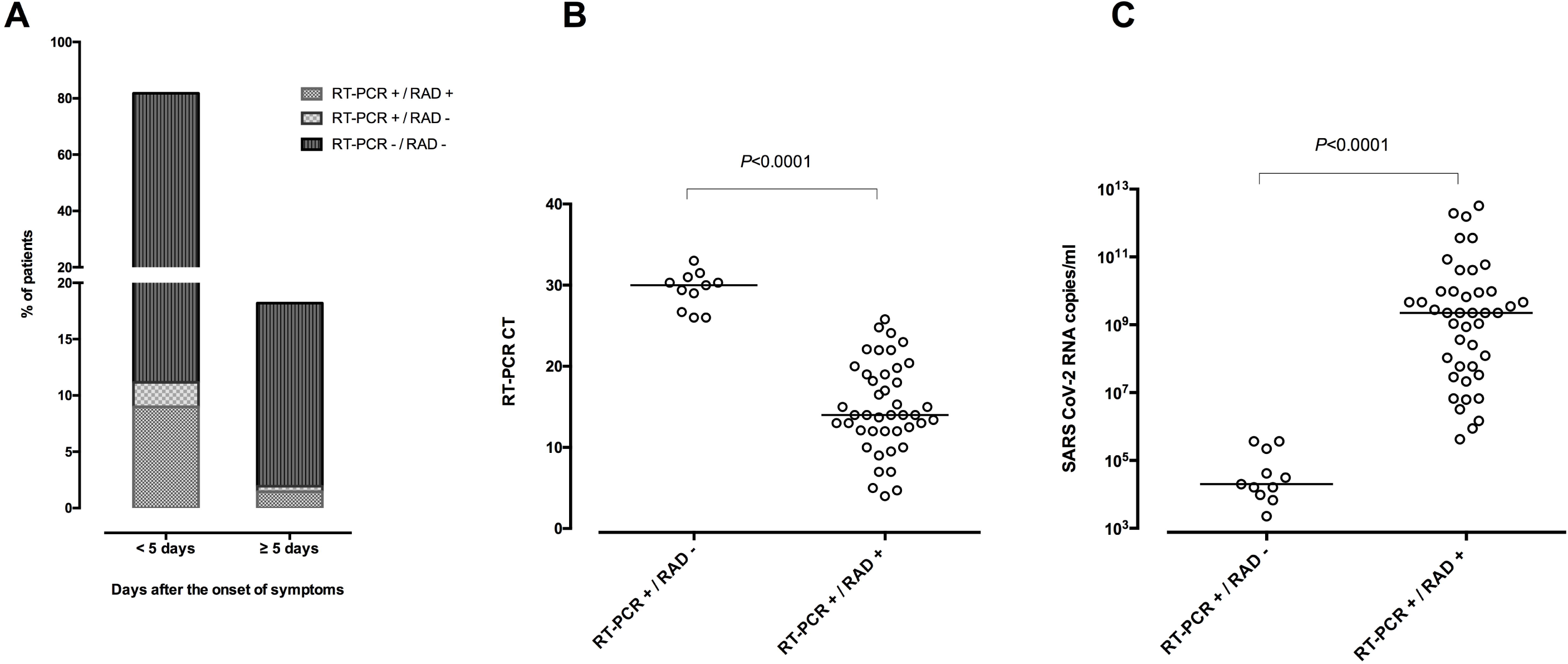
**(A)** Field performance of the Panbio™ COVID-19 AG Rapid Test Device (RAD) according to the time of testing since the onset of symptoms (< or ≥5 days) in a cohort of symptomatic patients with clinical suspicion of COVID-19 attended at primary healthcare centers. (B) RT-PCR C_T_ values in specimens testing either RAD + or RAD -. (C) SARS-CoV-2 RNA loads in specimens testing either RAD + or RAD-. Median and *P* values are shown.

The sensitivity was higher in adults (82.6%; 95% CI, 69.3-90.9%) than in pediatric patients (62.5%; 95% CI, 30.6-86.3%). The data are shown in Supplementary Table 2.

Overall RAD negative predictive value for an estimated prevalence of 5% and 10% was 99% (95% CI, 97.4-99.6%) and 97.9 (95% CI, 95.9-98.9), respectively.

RT-PCR C_T_ values and SARS-COV-2 RNA loads were significantly higher or lower (*P* <0.001), respectively, in RT-PCR+/RAD-than in RT-PCR+/RAD+ specimens (Figure 1B and 1C).

ROC curve analyses indicated that RT-PCR C_T_ <25 and SARS-CoV-2 RNA loads > 5.9 log_10_ copies/ml thresholds best discriminated between RT-PCR+/RAD+ and RT-PCR+/RAD-/ specimens, with a sensitivity and specificity of 100%

The time to sampling since the onset of symptoms did not differ (*P*=0.86) between RT-PCR+/RAD+ (median, 3 days; range 1-7 days) and RT-PCR+/RAD- (median 2 days; range, 1-6 days) patients.

Patients with fever, either with or without other accompanying symptoms, tested more frequently RT-PCR+/ RAD+ (*P*=0.02) than afebrile patients (Supplementary Table 3).

All 11 specimens yielding discordant RT-PCR/RAD results tested negative by culture, whereas SARS-CoV-2 could be recovered from 3 out of 3 specimens returning RT-PCR+/RAD+ results (C_T_: 4, 14 and 16).

## Discussion

Previous studies evaluating SARS-CoV-2 RAD tests either used retrieved specimens, which had been cryopreserved for a wide range of times, were conducted at central laboratories or both [3-7]. To our knowledge this is the first report on the performance of a RAD assay conducted at POC. As such, it may provide a realistic view of how implementation of RAD tests in clinical practice can contribute to the management and control of the COVID-19 pandemics. When compared to RT-PCR, the Panbio™ COVID-19 AG Rapid Test Device assay yielded an excellent specificity and a fairly good overall sensitivity (79.6%); the latter slightly improved when the time to testing was less than 5 days since the onset of symptoms (80.6%). This figure is less impressive than that claimed by the manufacturer (93%); however it is close (86.5%) to that reported by Linares and colleagues [3] in a mixed cohort including patients attended at the Emergency Department or primary healthcare centers and centralized testing at the hospital laboratory. Sensitivity of SARS-CoV-2 RAD assays has been reported to vary between 45%-97% [3-7]; yet, direct comparison between studies is hampered by relevant dissimilarities regarding clinical characteristics and age of patients, site of testing, type of specimen processed, and time to testing, among others.

Interestingly, the sensitivity was higher in adults (82.6%) than in pediatric patients (62.5%). Previous studies found no age-related differences in SARS-CoV-2 RNA load in the upper respiratory tract [8]. Although speculative, dating the accurate onset of symptoms could have been less reliable in children than in adults.

In a setting with an incidence of COVID-19 ranging between 5% and 10% such as ours at the time of the study, the RAD NPV was very good (99% and 97.9%, respectively).

There were 11 out of 54 RT-PCR positive specimens that tested negative by RAD. In line with previous reports [2-4], SARS-CoV-2 RNA load in RT-PCR+/ RAD+ specimens was significantly higher than that in RT-PCR+/ RAD-samples. In our setting, specimens with RT-PCR C_T_ >25 (equivalent to SARS-CoV-2 RNA loads < 5.9 log_10_ copies/ml) returned discordant RAD/RT-PCR results.

We did not observe a major impact of the time to testing on the likelihood of having a positive RAD result. On the other hand, fever was the only clinical feature reported more frequently in patients testing RT-PCR+/RAD+ than in those RT-PCR+/RAD-.

A relevant observation of our study was that SARS-CoV-2 could not be cultured from RT-PCR+/RAD-specimens. In line with that, Pekosz and colleagues [2], by using a highly sensitive cell culture system (VeroE6 TMPRSS2), found 1 out of 27 RAD-/culture + specimens. SARS-CoV-2 RNA load threshold associated with culture positivity herein (> 5.9 log_10_ copies/ml) was remarkably close to others previously published-around 10^6^ copies/ml-[2,9-11].

The main limitation of the current study is the relatively low number of cases in the series (13%); this, however, can be viewed as a strength, as this figure likely represents that found in many community settings worldwide where RAD testing is being increasingly used.

In summary, we found the Panbio™ COVID-19 AG Rapid Test Device to perform well as a POC for early diagnosis of COVID-19 in primary healthcare centers. More importantly, our data suggested that patients with RT-PCR-proven COVID-19 testing negative by RAD are unlikely to be infectious. Further studies are warranted to confirm this assumption. The inconsequentiality of false negative RAD results, from a public health perspective [12], would support the implementation of a laboratory diagnostic approach which excluded confirmatory RT-PCR testing for negative RAD tests in non-hospitalized patients, even when the pretest probability is high. This would certainly alleviate laboratory workloads while dealing with RT-PCR supply shortages.

## Supporting information

Signs and symptoms of patients with COVID-19 testing positive by RT-PCR and negative by the Panbio COVID-19 AG Rapid Test Device

RT-PCR and Rapid antigen test (RAD) results according to the age of patients (adults vs. pediatrics)

Demographic and clinical characteristics of patients testing SARS-CoV-2 RT-PCR positive Rapid antigen assay (RAD) positive or negative

## Data Availability

The authors confirm that the data supporting the findings of this study are available within the article [and/or] its supplementary materials.

## ACKNOWLEDGMENTS

We are grateful to Abbott Diagnostics for providing Panbio™ COVID-19 AG Rapid Test Device kits. We thank all personnel working at Microbiology Service of Clinic University Hospital for their unwavering commitment in the fight against COVID-19. We also thank María José Beltrán, Pilar Botija and Ana Sanmartín for assistance in the organization of RAD testing in primary healthcare centers.

## AUTHOR’S CONTRIBUTIONS

EA, IT, FB, DH, MM, SP, LF, AV, CSdA, JP and JC: Methodology (RT-PCR and RAD) and validation of data; EM and MA F-F: Methodology (cell culture) and validation of data. EA, IT: formal analysis. DN: Conceptualization, supervision, writing the original draft. All authors reviewed the original draft.

