## Supplementary material for "Field evaluation of a rapid antigen test (Panbio™ COVID-19 Ag Rapid Test Device) for the diagnosis of COVID-19 in primary healthcare centers": Signs and symptoms of patients with COVID-19 testing positive by RT-PCR and negative by the Panbio COVID-19 AG Rapid Test Device

| **Supplementary TABLE 1. Signs and symptoms of patients with COVID-19 testing positive by RT-PCR and negative by the Panbio™ COVID-19 AG Rapid Test Device** | | | | | | | | | | | | |
| --- | --- | --- | --- | --- | --- | --- | --- | --- | --- | --- | --- | --- |
| **Day after the onset of signs and symptoms** | **Age** | **RT-PCR C_T_/log_10_ RNA copies/ml** | **Dry cough** | **Fever** | **Odynophagia** | **Cephalea** | **Anosmia** | **Ageusia** | **Diarrhoea** | **Dyspnea** | **Exanthema** | **Myalgia** |
| 3 | 25 | 31/3.9 | X |  |  |  |  |  |  |  |  |  |
| 1 | 27 | 30.3/4.2 |  |  |  |  | X | X |  |  |  |  |
| 2 | 5 | 30/4.3 |  | X |  |  |  |  |  |  |  |  |
| 1 | 6 | 30.3/4.2 | X | X | X |  |  |  |  |  |  |  |
| 2 | 38 | 26.7/5.3 | X |  |  |  |  |  |  |  |  |  |
| 5 | 20 | 26/5.6 |  |  |  |  |  |  |  |  | X |  |
| 6 | 13 | 33/3.3 | X | X |  | X |  |  | X |  |  |  |
| 2 | 70 | 26/6.6 | X |  | X |  |  |  |  |  |  |  |
| 3 | 39 | 29/4.6 |  |  |  |  | X |  |  |  |  | X |
| 2 | 42 | 31.5/3.8 | X | X |  |  |  |  |  |  |  | X |
| 3 | 69 | 29.4/4.5 |  |  | X |  |  |  |  |  |  |  |
