## Supplementary material for "Field evaluation of a rapid antigen test (Panbio™ COVID-19 Ag Rapid Test Device) for the diagnosis of COVID-19 in primary healthcare centers": RT-PCR and Rapid antigen test (RAD) results according to the age of patients (adults vs. pediatrics)

| **Supplementary TABLE 2. RT-PCR and Rapid antigen test (RAD) results according to the age of patients (adults vs. pediatrics)** | | | |
| --- | --- | --- | --- |
| Patient population | RT-PCR result | RAD positive  no. of specimens/median RT-PCR cycle threshold (range) | RAD negative  no. of specimens/ median RT-PCR cycle threshold (range) |
| Adult (>16 years old) | Positive | 38/14.5 (4-25.8) | 8/29.2 (26-31.5) |
|  | Negative | 0/- | 281/- |
| Pediatric (≤16 years old) | Positive | 5/10 (4.7-14) | 3/30.3 (30-33) |
|  | Negative | 0/- | 77/- |
