## Supplementary material for "Field evaluation of a rapid antigen test (Panbio™ COVID-19 Ag Rapid Test Device) for the diagnosis of COVID-19 in primary healthcare centers": Demographic and clinical characteristics of patients testing SARS-CoV-2 RT-PCR positive Rapid antigen assay (RAD) positive or negative

| **Supplementary TABLE 3. Demographic and clinical characteristics of patients testing SARS-CoV-2 RT-PCR positive Rapid antigen assay (RAD) positive or negative** | | |
| --- | --- | --- |
|  | RAD positive (n=43) | RAD negative (n=11) |
| Age (median, range) | 29 (4-77) | 27 (5-70) |
| Sex (Male/Female) | 21 / 22 | 4 / 7 |
| Days after the onset of symptoms (median, range) | 3 (1-7) | 2 (1-6) |
| Fever; no. (%) | 33 (76,7) | 4 (36,4) |
| Cough; no. (%) | 14 (32,6) | 6 (54,5) |
| Odynophagia; n (%) | 8 (18,6) | 3 (27,3) |
| Cephalea; n (%) | 10 (23,3) | 1 (9,1) |
| Anosmia; n (%) | 7 (16,3) | 2 (18,2) |
| Ageusia; n (%) | 5 (11,6) | 2 (18,2) |
| Diarrhoea; n (%) | 11 (25,6) | 1 (9,1) |
| Dyspnoea; n (%) | 3 (7,0) | 0 (0,0) |
| Rash; n (%) | 0 (0,0) | 1 (9,1) |
| Myalgia; n (%) | 16 (37,2) | 2 (18,2) |
